# Maladaptive Self-Focused Attention and Default Mode Network Connectivity: A Transdiagnostic Investigation Across Social Anxiety and Body Dysmorphic Disorders

**DOI:** 10.1101/2021.05.25.21257688

**Authors:** Angela Fang, Bengi Baran, Clare C. Beatty, Jennifer Mosley, Jamie D. Feusner, K. Luan Phan, Sabine Wilhelm, Dara S. Manoach

## Abstract

**Background:** Maladaptive self-focused attention (SFA) is a bias toward internal thoughts, feelings, and physical states. Despite its role as a core maintaining factor of symptoms in cognitive theories of social anxiety and body dysmorphic disorders, studies have not examined its neural basis. We hypothesized that maladaptive SFA would be associated with hyperconnectivity in the default mode network (DMN) in self-focused patients with these disorders.

**Methods:** Thirty patients and 28 healthy individuals were eligible and scanned. Eligibility was determined by scoring greater than 1SD or below 1SD of the Public Self-Consciousness Scale normative mean, respectively, for each group. Fifteen patients had primary social anxiety disorder and 15 had primary body dysmorphic disorder. Seed-to-voxel functional connectivity was computed using a DMN posterior cingulate cortex (PCC) seed.

**Results:** Patients (regardless of diagnosis) showed reduced functional connectivity of the PCC with several brain regions, including the bilateral superior parietal lobule (SPL), bilateral insula, cingulate cortex, and postcentral gyrus, compared to controls. PCC-SPL connectivity was inversely correlated with maladaptive SFA in patients but was not associated with social anxiety or body dysmorphic symptom severity, depression severity, or rumination. There was no evidence of increased functional connectivity within the DMN in patients compared to controls.

**Conclusions:** As the SPL is part of the dorsal attention network, which is typically activated during tasks requiring externally-oriented attention, abnormal PCC-SPL connectivity in patients may reflect difficulty shifting between internal versus external attention, and may represent a transdiagnostic neural marker of maladaptive SFA that could be targeted in interventions.

**Clinical Trials Registration:** ClinicalTrials.gov Identifier: NCT02808702 https://clinicaltrials.gov/ct2/show/NCT02808702

## Introduction

Self-focused attention (SFA) is a form of self-referential thought that falls along a dimension, with an adaptive “experiential” mode of processing on one end, and a maladaptive “analytical” mode on the other (1). Maladaptive SFA is a dispositional tendency to reflect excessively about one’s internal thoughts, feelings, beliefs, and physical states that interferes with attention to the external environment (2). Neurocognitive models of psychiatric disorders propose that attentional biases, such as maladaptive SFA, lead to negatively-biased self-referential thoughts, which increase negative mood (e.g., anxiety, depression), and maladaptive behaviors (e.g., avoidance, safety seeking behaviors) (2-4). These attentional biases may reflect disrupted top-down attentional control or bottom-up attentional orienting mechanisms (5-6). Transdiagnostically, maladaptive SFA conceptually overlaps with ruminations in depression (7), obsessions in obsessive-compulsive disorder (8), worry in generalized anxiety disorder (9), anxiety sensitivity in panic disorder (10), and somatic preoccupations in eating disorders (11). As research on SFA has been largely disorder-specific, studies have not tested whether maladaptive SFA extends across disorders, and the neural correlates of maladaptive SFA remain unknown. In this study, we tested whether maladaptive SFA would be associated with hyperconnectivity within the Default Mode Network across two self-focused psychiatric disorders.

Functional neuroimaging studies suggest that SFA is associated with activity in a large-scale, intrinsic brain network that mediates internal mentation: the Default Mode Network (DMN) (12-14). The DMN comprises several brain regions that form an integrated system for self-related cognition, including the medial prefrontal cortex (mPFC) and portions of the anterior cingulate cortex (ACC), precuneus/posterior cingulate cortex (PCu/PCC), angular gyrus, lateral parietal cortex, and medial temporal cortex (15-19). Among these regions, the mPFC and the PCC display consistent patterns of activity across a variety of internally-directed tasks and processes (20-22). Brain regions within the DMN are commonly deactivated during tasks requiring external attention, a phenomenon known as task deactivation or task suppression (23), which suggests that orienting attention between internal and external environments requires balance between the DMN and other networks (24).

A growing body of literature using task and resting state fMRI in “self-focused” clinical and non-clinical populations implicate abnormalities in the DMN (25-27). For example, one study found that maladaptive SFA was associated with increased activation in the mPFC, temporo-parietal junction (TPJ), and temporal pole (TP), in a sample of individuals with high socially anxious symptoms (28). A study in body dysmorphic disorder found reduced DMN deactivation in the dorsal mPFC during a house-viewing task, which may be associated with a reduced ability to disengage the DMN during the task due to self-referential thinking (29).

Similarly, impaired deactivation of the DMN during error processing has been demonstrated in a study in obsessive-compulsive disorder (30), which may be suggestive of persistent self-referential processing after making errors. Studies examining resting state functional connectivity in patients with social anxiety disorder have shown both increased and decreased functional connectivity within the DMN, in patients compared to controls (31-34), as well as one study reporting no group differences in a small sample of drug-naïve, non-comorbid individuals (35). Although social anxiety disorder is associated with high levels of maladaptive SFA (36), these studies did not explicitly link DMN connectivity with SFA. Furthermore, inconsistent findings may be due to methodological differences in resting state protocols and data processing procedures (especially in addressing motion-related confounds), as well as sample differences in symptoms and severity in psychiatric disorders. More research is needed to examine the extent to which maladaptive SFA is associated with DMN disruptions in clinical populations.

We addressed limitations of the prior literature by employing a novel sampling strategy and taking rigorous steps to account for motion-related confounds in functional connectivity. First, we recruited individuals based on self-reported levels of maladaptive SFA: patients with either social anxiety disorder (SAD) or body dysmorphic disorder (BDD) who scored above a cutoff on the Public Self-Consciousness Scale, a measure of trait SFA (37), and healthy individuals who scored below the cutoff. We selected these two related disorders on the anxiety and obsessive-compulsive spectrum for the following reasons: (1) maladaptive SFA is proposed to be a major maintaining factor of symptoms in both disorders (3-4); (2) both disorders share a core fear of negative evaluation by others, which activates maladaptive SFA and results in social avoidance (38); and (3) a majority of the literature on SFA has focused on depression.

These studies tend to emphasize the ruminative aspects of SFA in depression, as compared to the social presentation and self-consciousness aspects of SFA more relevant in anxiety and obsessive-compulsive related disorders (1,3,13). Second, we rigorously controlled for the confounding effects of head motion in functional connectivity studies by training participants to remain still during mock scan sessions and by procedures implemented during both data acquisition and postprocessing (described below). We hypothesized that patients with maladaptive SFA (across both SAD and BDD diagnoses) would display greater resting state functional connectivity within the DMN, as compared to controls. We also predicted that connectivity within the DMN would correlate more strongly with self-reported trait maladaptive SFA (as compared to related constructs, such as depression severity and rumination) across both disorders, given the transdiagnostic nature of maladaptive SFA.

## Methods and Materials

### Participants

Patients were treatment-seeking individuals recruited from outpatient psychiatry specialty clinics at the Massachusetts General Hospital and healthy individuals were recruited from the community through flyers and online advertisements. A total of 89 potentially eligible participants, who met prescreening criteria, consented to the study. Participants were recruited on the basis of their self-reported levels of trait maladaptive SFA, as measured by the Public Self-Consciousness Scale-Revised (SCS-R) (37). Included patients scored _≥_18 on this measure, whereas healthy controls were required to score ≤9, which reflects +/-1SD of the normative mean, respectively. There are 7 items in this measure, scored on a 0 (not at all like me) to 3 (a lot like me) Likert scale. Example items include: *I care a lot about how I present myself to others, I’m self-conscious about the way I look, and I usually worry about making a good impression*. The Public SCS-R was given during pre-screening prior to enrollment, as well as during an in-person screening evaluation to confirm trait levels of SFA. Patients also had to meet cutoff scores reflecting at least moderate levels of SAD or BDD symptoms (≥50 on the Liebowitz Social Anxiety Scale [LSAS] (39) for those with primary SAD, and ≥20 on the Yale-Brown Obsessive-Compulsive Scale Modified for BDD [BDD-YBOCS] (40) for those with primary BDD). Diagnoses were evaluated using the SCID-5 (41). Low-self-focused healthy individuals had no lifetime history of psychiatric disorders. Exclusion criteria for all participants included: (1) MR contraindications (e.g., claustrophobia, metal in body, pregnancy), (2) history of head injury, neurological disorder, or neurosurgical procedure, (3) active suicidal or homicidal ideation, (4) lifetime manic, hypomanic, or psychotic episode, (5) past year alcohol or substance dependence, or (6) current or past cognitive behavioral therapy or formal mindfulness/meditation training, defined by >10 sessions.

After the screening evaluation, 25 were ineligible for the following reasons: no longer met criteria on the SCS-R score (n=4 patients, n=5 controls), not meeting criteria for primary SAD or primary BDD (n=9 patients), current or past cognitive behavioral therapy or formal meditation (n=4 patients), trying to get pregnant (n=1 patient), history of alcohol use disorder (n=1 control), and inadequate match with a clinical patient (n=1 control). An additional 6 participants withdrew prior to completing the fMRI scan due to time commitment (n=2 patients, n=1 control), lost to follow-up (n=2 controls), and moved out of state (n=1 control). This left a final sample of 30 patients (15 with a primary diagnosis of SAD, 15 with a primary diagnosis of BDD) and 28 age-, sex-, and education-matched healthy individuals.

### Procedures

Participants attended an initial evaluation, which involved a comprehensive diagnostic interview using the SCID-5, clinician-administered symptom assessments (e.g., LSAS, BDD-YBOCS, and Brown Assessment of Beliefs Scale [BABS] (42) to assess levels of insight), and self-report measures of depression severity using the Beck Depression Inventory-II (BDI-II) (43), rumination using the Ruminative Responses Scale (RRS) (44), handedness using the Edinburgh Handedness Scale (45), and estimated verbal IQ using the Wide Range Achievement Test-4 (WRAT-4) (46). Eligible participants returned for an MRI scan between 7-10 days of the initial evaluation. The study was approved by the Partners Human Research Committee and all participants provided written informed consent.

### MRI Acquisition

Images were acquired with a 3T Siemens Prisma scanner (Siemens Medical Systems, Iselin, NJ) equipped for echo planar imaging and a 64-channel head coil. Participants who had never undergone MRI scans completed a mock scan on a separate day or just prior to their actual scan to acclimate them to the scanning environment and to train them to remain still, with the exception of 1 patient with body dysmorphic disorder and 2 healthy controls. During scanning, Head movements were restricted using foam cushions. A high-resolution anatomical scan was acquired using a T1-weighted 3D multiecho magnetization-prepared rf-spoiled rapid gradient-echo MEMPRAGE sequence with EPI based volumetric navigators for prospective motion correction and selective reacquisition (47) (TR=2530ms, flip angle=7°, TEs=1.7ms/3.6ms/5.4ms/7.3ms, iPAT=2, FOV=256mm, 176 in-plane sagittal slices; voxel size=1mm^3^ isotropic, scan duration 5m 34s). Two resting state functional connectivity MRI scans were acquired with a T2*-weighted gradient echo sequence for blood oxygen level dependent (BOLD) contrast (TR=2000ms, flip angle=85°, TE=30ms, FOV=205mm, 32 continuous horizontal slices parallel to the intercommissural plane, voxel size=3.2×3.2×3.3mm, interleaved, scan duration 6 min 6s). The resting state sequences included prospective acquisition correction (PACE) for head motion to adjust slice position and orientation during image acquisition (48). Participants were instructed to keep their eyes open for the duration of the scan.

### MRI Preprocessing and Data Quality

Automated preprocessing steps were performed using the CONN toolbox version 17 (49), which employs routines from the Statistical Parametric Mapping software (SPM8; Wellcome Trust Centre for Neuroimaging, London, UK). The first four volumes of each resting state run were discarded to allow for T1 equilibration. Images were segmented into gray matter, white matter, and cerebrospinal fluid masks, corrected for slice timing, and spatially realigned to the reference image. The volumes were then normalized to the MNI template provided in SPM8, resampled to 2mm voxels, and spatially smoothed using a Gaussian kernel with a full width at half maximum of 8mm. A temporal band-pass filter of .008-.09 Hz was applied to the time series. Nuisance variables (white matter, cerebrospinal fluid, movement parameters) were addressed using the anatomical CompCor method (50), which uses a component base noise reduction method, rather than global signal regression (51), to reduce physiological and other noise. Artifactual volumes were identified using the Artifact Detection Tools (ART; http://www.nitrc.org/projects/artifact_detect/) and defined as volumes with > 0.9 mm head displacement in the x, y or z directions or global mean intensity more than 5 SDs above the entire scan. Residual head motion parameters (rotation and translations in x, y, and z directions and their first-order temporal derivatives, calculated using Power et al.’s (52) measure of framewise displacement) and artifactual volumes (flagged by ART) were regressed out (i.e. scrubbed) in the model. There were no group differences in residual head motion, number of artifactual volumes, or proportion of scrubbed volumes to total number of volumes, in either resting state run (see Table 1).

**Table 1.** Sample Characteristics. WRAT-4 = Wide Range Achievement Test-4. Score reflects word reading subtest standard score. Head motion based on Power et al.’s (52) measure of framewise displacement. BDI-II = Beck Depression Inventory-II. LSAS = Liebowitz Social Anxiety Scale. BDD-YBOCS = Yale-Brown Obsessive-Compulsive Scale Modified for BDD. BABS = Brown Assessment of Beliefs Scale. Public SCS-R = Public Subscale of Self-Consciousness Scale-Revised. RRS = Ruminative Responses Scale. ^a^t-statistics reflect differences between social anxiety disorder (SAD) and body dysmorphic disorder (BDD) groups.

| **Demographic Characteristics** | **Clinical (n=30)**<br>Mean ± SD / n (%) | **Control (n=28)**<br>Mean ± SD / n (%) | **t / $\chi^2$** | **p** |
| --- | --- | --- | --- | --- |
| Age | 25.43 ± 3.76 | 25.71 ± 3.89 | 0.28 | 0.78 |
| WRAT-4 | 71.50 ± 22.57 | 74.04 ± 19.08 | 0.46 | 0.65 |
| Sex |  |  | 0.20 | 0.65 |
| Female | 23 (76.7) | 20 (71.4) |  |  |
| Male | 7 (23.3) | 8 (28.6) |  |  |
| Race |  |  | 1.65 | 0.65 |
| Caucasian | 19 (63.3) | 18 (64.3) |  |  |
| Asian | 8 (26.7) | 5 (17.9) |  |  |
| Black/African American | 2 (6.7) | 2 (7.1) |  |  |
| More than one race | 1 (3.3) | 3 (10.7) |  |  |
| Ethnicity |  |  | 0.09 | 0.76 |
| Non-Hispanic or Latino | 26 (86.7) | 25 (89.3) |  |  |
| Hispanic or Latino | 4 (13.3) | 3 (10.7) |  |  |
| Unspecified | 0 (0.0) | 0 (0.0) |  |  |
| Highest Level of Education |  |  | 2.96 | 0.40 |
| Some College | 3 (10.0) | 0 (0.0) |  |  |
| College Graduate | 17 (56.7) | 18 (64.3) |  |  |
| Some Postgraduate | 1 (3.3) | 1 (3.6) |  |  |
| Postgraduate/Professional Degree | 9 (30.0) | 9 (32.1) |  |  |
| Handedness | 77.83 (33.21) | 86.43 (17.84) | -1.22 | 0.23 |
| Head motion |  |  |  |  |
| Resting state run 1 | 0.16 ± 0.11 | 0.15 ± 0.05 | -0.19 | 0.85 |
| Resting state run 2 | 0.18 ± 0.09 | 0.20 ± 0.09 | 0.95 | 0.35 |
| Number of artifactual volumes (% scrubbed volumes of total volumes) |  |  |  |  |
| Resting state run 1 | 9.57 (5.41%) | 8.00 (4.52%) | -0.71 | 0.48 |
| Resting state run 2 | 7.63 (4.31%) | 6.92 (3.91%) | -0.60 | 0.55 |

| Clinical Characteristics | BDD (n=15)<br>Mean ± SD<br>/ n (%) | SAD (n=15)<br>Mean ± SD<br>/ n (%) | | t<sup>a</sup> | p |
| --- | --- | --- | --- | --- | --- |
| Age of onset (years) | 14.20 ± 2.62 | 13.20 ± 3.21 | --- | 0.93 | 0.36 |
| BDI-II | 15.53 ± 12.51 | 17.00 ± 14.65 | 0.39 (0.69) | 0.30 | 0.77 |
| LSAS | --- | 80.27 ± 16.04 | --- | --- | --- |
| BDD-YBOCS | 30.20 ± 6.57 | --- | --- | --- | --- |
| BABS | 14.80 ± 4.36 | 14.33 ± 2.90 | --- | 0.35 | 0.73 |
| Public SCS-R | 20.07 ± 1.10 | 20.07 ± 1.28 | 4.57 (2.43) | 0.00 | 1.00 |
| RRS | 53.13 ± 13.97 | 53.33 ± 16.42 | 28.63 (8.01) | 0.04 | 0.97 |

### Functional Connectivity Analyses

Seed-to-voxel functional connectivity analyses of the DMN were conducted using the canonical DMN seed, the posterior cingulate cortex (PCC), provided by the CONN toolbox (49). First level functional connectivity maps were generated by extracting the average BOLD time series from the PCC seed and correlating it with every gray matter voxel in the whole brain. Correlation values were then transformed using Fisher’s z transformation to yield a map for each resting state run in which the value at each voxel represented connectivity with the PCC seed. Two resting state runs for each subject were averaged. We examined group differences in DMN connectivity with t-tests at every voxel. Whole brain correction for multiple comparisons was applied using a voxel level uncorrected threshold of *p*<.001 and a false discovery rate (FDR)-corrected cluster threshold of *p*<.05.

In the patient group, we conducted correlation analyses to test the relations between DMN connectivity in regions that significantly differed between groups in the whole brain and trait maladaptive SFA (Public SCS-R), and other clinical variables (depression severity, rumination, social anxiety severity, and body dysmorphic severity). To determine whether correlations with trait maladaptive SFA were significantly stronger than correlations with other clinical variables, we used Steiger’s (53-54) calculation for testing the equality of two correlation coefficients measured on the same individuals. As our *a priori* hypotheses were that DMN connectivity measures would show a stronger association with trait maladaptive SFA compared to other clinical variables, these analyses were tested at *p*<.05.

## Results

### Sample characteristics

As shown in Table 1, there were no differences between patients and controls in age, sex, race, ethnicity, education, handedness, and estimated verbal IQ. Five patients were taking the following medications at a stable dose for at least two months prior to study enrollment: escitalopram (n=1, SAD), citalopram (n=3, BDD), and amphetamine (n=1, BDD). Full sample details regarding psychiatric comorbidities are described in supplementary materials (Table S1).

### Abnormal DMN functional connectivity in self-focused patients with SAD and BDD

Contrary to our hypothesis, no regions showed significantly greater functional connectivity within the DMN in patients versus controls. Compared with healthy controls, patients exhibited reduced functional connectivity between the PCC and a set of cortical regions, including the right cingulate cortex, bilateral superior parietal lobule, left postcentral gyrus, and bilateral insula (Table 2; Figure 1A). None of these regions were within the DMN. These group differences were driven by weak or slightly positive connectivity in controls and slightly negative connectivity in patients, in all regions, except the right anterior insula, which showed negative connectivity in both groups (Figure 2 shows unthresholded statistical maps of PCC functional connectivity).

**Table 2.**
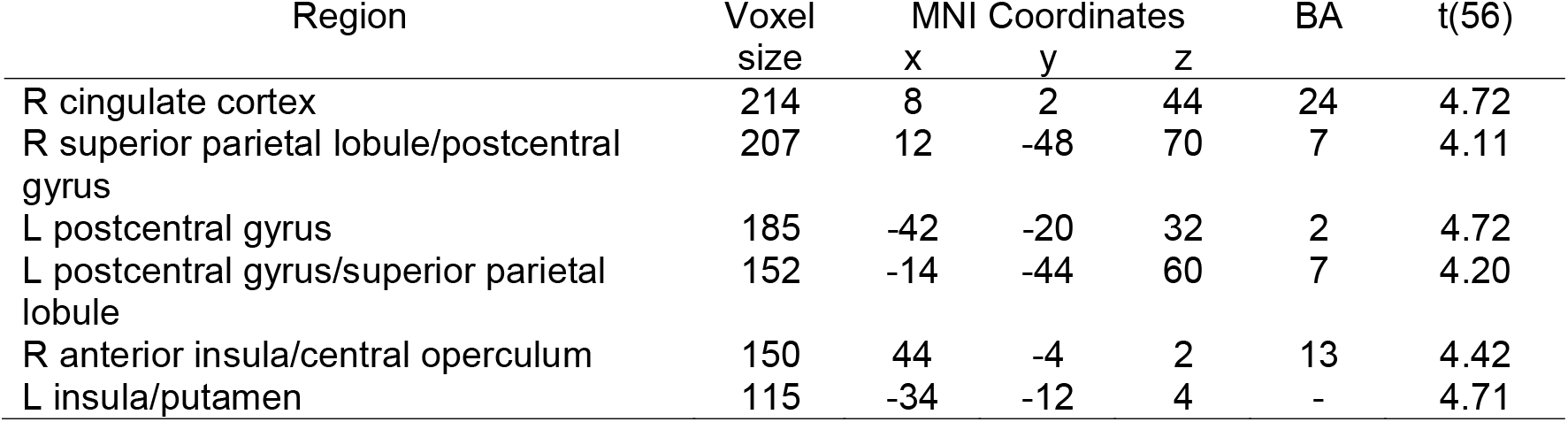
Clusters showing significant group differences in functional connectivity with the posterior cingulate cortex (PCC) All reported clusters are significant at p_FDR-corrected_ <.05, two-sided, based on whole-brain correction. There were no clusters in which patients showed significantly greater functional connectivity than controls. Montreal Neurological Institute (MNI) coordinates and t-statistics are given for peak voxel location.

| Region | Voxel<br>size | MNI Coordinates |  |  | BA | t(56) |
| --- | --- | --- | --- | --- | --- | --- |
|  |  | x | y | z |  |  |
| R cingulate cortex | 214 | 8 | 2 | 44 | 24 | 4.72 |
| R superior parietal lobule/postcentral<br>gyrus | 207 | 12 | -48 | 70 | 7 | 4.11 |
| L postcentral gyrus | 185 | -42 | -20 | 32 | 2 | 4.72 |
| L postcentral gyrus/superior parietal<br>lobule | 152 | -14 | -44 | 60 | 7 | 4.20 |
| R anterior insula/central operculum | 150 | 44 | -4 | 2 | 13 | 4.42 |
| L insula/putamen | 115 | -34 | -12 | 4 | - | 4.71 |

**Figure 1.**
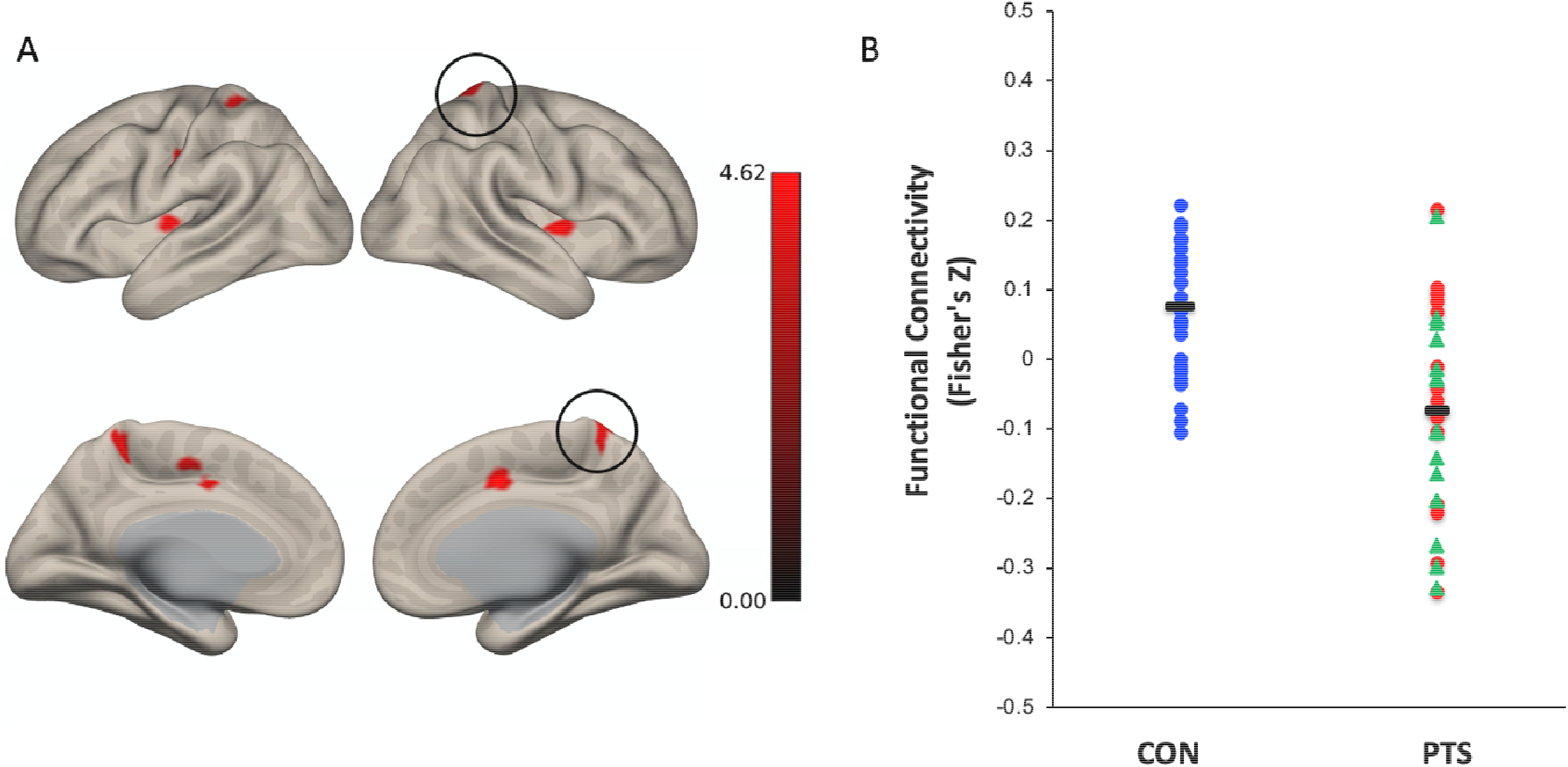
Group differences in DMN connectivity. (A) Statistical map of group differences in lateral and medial views of cortical surface at p_FDR-corrected_ <.05. Greater connectivity in controls is depicted in red. There were no regions of significantly greater connectivity in patients. PCC-right SPL connectivity is circled. (B) Distribution of connectivity between PCC seed and right SPL cluster is shown separately in each group. Black bars represent group means. (C) Scatterplot of PCC-right SPL connectivity in patients with trait SFA scores on Public SCS-R measure. Green triangles represent patients with SAD. Red circles represent patients with BDD.

**Figure 2.**
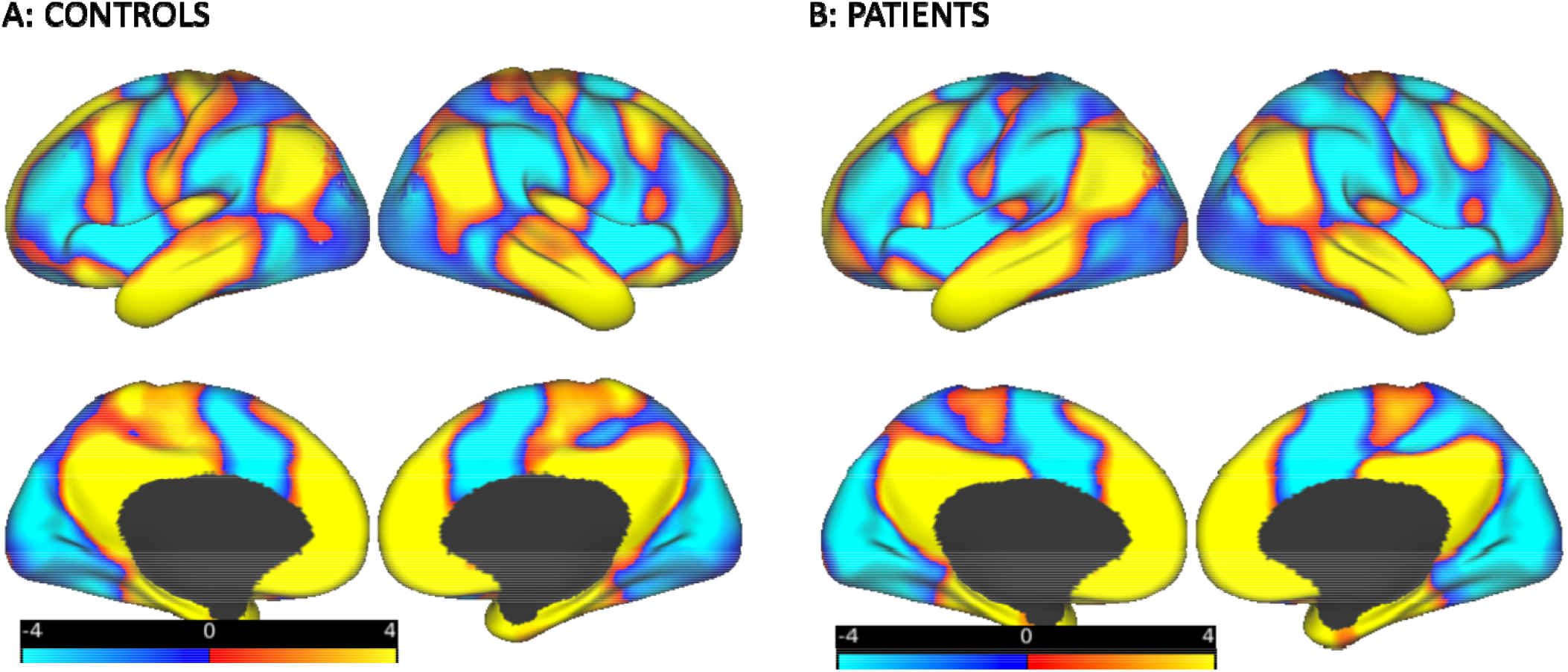
Unthresholded statistical maps of PCC functional connectivity displayed on a template brain. (A) Healthy control group. (B) Patients with SAD and BDD. Positive connectivity is displayed in warm colors; negative connectivity is displayed in blue. Units on the color key reflect t-values.

We conducted the same analysis while controlling for concurrent medications by covarying medication status. This resulted in a similar pattern of results, but only at a voxel-level threshold of *p*<.005 (cluster-level FDR-corrected threshold of *p*<.05; Supplementary Material Table S3).

### Associations between DMN functional connectivity and maladaptive SFA

Connectivity measures within the six regions showing group differences were extracted for individual subjects and correlated with Public SCS-R scores, BDI-II, RRS, LSAS, and BDD-YBOCS scores. There were two significant inverse correlations between PCC-bilateral superior parietal lobule connectivity and Public SCS-R scores (right superior parietal lobule: *r* = -.573, *p* = .001; left superior parietal lobule: *r* = -.530, *p* = .003), reflecting that greater SFA was associated with reduced connectivity of the PCC with the superior parietal lobule (Table 3). PCC-bilateral superior parietal lobule connectivity did not correlate significantly with either depression severity, rumination, social anxiety or body dysmorphic symptom severity. Only for right PCC-superior parietal lobule connectivity were correlations with Public SCS-R scores significantly different from correlations with BDI-II (*p*=.05), RRS (*p*=.05), and BDD-YBOCS (*p*=.01), but not LSAS scores (*p*=.37). The correlations for left PCC-left superior parietal lobule connectivity with Public SCS-R scores did not significantly differ from the other clinical measures (all *p*’s>.14). A full correlation matrix of connectivity and clinical measures in patients and controls are reported in Table S2.

**Table 3.** Correlations Between PCC Connectivity and Clinical Measures in Patients Only. Table 3. PCC = posterior cingulate cortex. CC = cingulate cortex. SPL = superior parietal lobule. PCG = postcentral gyrus. INS = insula. LSAS = Liebowitz Social Anxiety Scale. BDD-YBOCS = Yale-Brown Obsessive-Compulsive Scale Modified for BDD. BDI-II = Beck Depression Inventory-II. RRS = Ruminative Responses Scale.

|  | Public SCS-R | LSAS | BDD-YBOCS | BDI-II | RRS |
| --- | --- | --- | --- | --- | --- |
| 1. PCC-right CC | -.198 | -.398 | -.378 | -.178 | -.232 |
| 2. PCC-right SPL | -.573*** | -.417 | -.049 | -.150 | -.133 |
| 3. PCC-left PCG | .034 | -.552* | -.439 | -.405* | -.433* |
| 4. PCC-left SPL | -.530** | -.464 | -.345 | -.265 | -.198 |
| 5. PCC-right INS | -.017 | -.116 | -.615* | -.280 | -.402* |
| 6. PCC-left INS | -.018 | -.127 | -.182 | -.242 | -.259 |

In patients, PCC connectivity with the left postcentral gyrus was significantly correlated with depression severity, rumination, and social anxiety severity, and PCC connectivity with the right anterior insula was significantly correlated with rumination and body dysmorphic severity (all *p*’s<.05). No other DMN connectivity measures were significantly correlated with clinical measures in patients. In controls, none of the connectivity measures were significantly associated with maladaptive SFA, depression severity, or rumination.

To examine whether DMN connectivity differences between patients and controls were driven by either the SAD or BDD group, we compared the six connectivity measures in simple post-hoc comparisons between SAD vs controls, BDD vs controls, and SAD vs BDD. Our post-hoc tests showed that connectivity differences between individuals with SAD vs controls remained (all *p*’s≤.001), and connectivity differences between BDD vs controls (all *p*’s≤.005) remained, in the same direction of our original findings. There were no significant differences in connectivity between SAD vs BDD (*p*’s ranging between .21 to .87).

## Discussion

Contrary to our hypothesis, we did not find evidence of greater resting state functional connectivity in the DMN in patients with SAD or BDD with maladaptive SFA. Our main finding was that patients, relative to controls, exhibited abnormally reduced functional connectivity between the PCC, a key component of the DMN, and brain regions that are typically activated during cognitive task execution, specifically dorsal cingulate cortex, bilateral superior parietal lobule, bilateral insula, and postcentral gyrus, which reflect certain anatomical components of the dorsal attention network (24,55) and salience network (56-57). These connectivity differences were not driven by one of the two patient groups, as the same pattern of differences emerged within both groups and there were no significant differences between groups. Of all the connectivity measures that showed group differences, only PCC connectivity with bilateral superior parietal lobule was significantly associated with self-reported maladaptive SFA in patients, in an inverse direction, such that greater maladaptive SFA was associated with reduced PCC-bilateral superior parietal lobule connectivity. Our results suggest that maladaptive SFA may reflect a failure to modulate the balance of attention between internal and external milieus in both disorders.

There were several regions in our results that belong to the “task positive network”, which typically includes the intraparietal sulcus, inferior parietal lobule, frontal eye fields, dorsolateral prefrontal cortex, middle temporal regions, insula, and supplementary motor area (24). Our results suggest that abnormal interactions between the DMN and such task positive regions as the superior parietal lobule, postcentral gyrus, and anterior insula may reflect difficulties shifting between these networks. The superior parietal lobule is considered a component of the dorsal attention network (DAN), together with the frontal eye fields and intraparietal sulcus, which become activated during directed attention (55). In our study, reduced PCC-superior parietal lobule connectivity may reflect abnormal DMN-DAN connectivity, as an index of the biased attention toward internal processing (at the detriment of external processing) that is characteristic of maladaptive SFA, which is consistent with two recent studies in pediatric obsessive-compulsive disorder showing similar DMN-DAN functional connectivity disruptions (58-59). However, our finding contrasts with prior literature showing that the DMN and DAN are typically anticorrelated networks, as we found weak or slightly positive DMN-DAN connectivity in controls and negative DMN-DAN connectivity in patients (as shown in Figure 1B). This may point to the complex and dynamic nature of voluntary attentional orienting that is not captured by our functional connectivity measure (which takes the correlation between the average time series of two brain regions) and may be better assessed using time-varying dynamic connectivity, as discussed by Maillet and colleagues (60). For example, it is possible that the content of thoughts differed between patients and controls during the resting state scan, which led to varying amounts of sustained attention toward internal thoughts. Another possibility is that our sampling strategy recruited especially “low self-focused” healthy controls, and it remains unclear whether such participants would perform differently on cognitive tasks or display different connectivity patterns.

There was some mixed evidence in support of PCC connectivity with the superior parietal lobule as specifically reflecting maladaptive SFA across patients with SAD and BDD. In subsequent tests comparing associations between connectivity and clinical variables, there was some evidence supporting its specific association with maladaptive SFA, which was stronger and significantly different from its association with depression severity, rumination, and body dysmorphic severity. However, its association with maladaptive SFA and social anxiety severity did not differ significantly. This is not surprising given that maladaptive SFA is typically highly correlated with social anxiety severity (36), and had a correlation of *r*=.378 in our sample. In fact, we expected maladaptive SFA to be more highly correlated with both SAD and BDD severity, given that it is such a prominent feature of the two conditions. The lower strength of the associations that we found may be due to restricted range on the upper tail of the Public SCS-R measure. It is also noteworthy that this measure captures the social self-presentation concerns that are more characteristic of SFA in SAD and BDD, compared to SFA in depression, which is more ruminative in nature (1,7). This explains why PCC-right superior parietal lobule connectivity may specifically index maladaptive SFA in SAD, but not in depression. Interestingly, our left-sided measure of PCC-superior parietal lobule connectivity was not found to be specifically associated with maladaptive SFA, relative to other clinical variables. This contrasts with evidence supporting a largely bilateral dorsal attention system (55), which is consistent with a study in obsessive-compulsive disorder showing abnormalities in DMN subsystem functional connectivity with bilateral superior parietal lobule (61), but inconsistent with a study in SAD showing left lateralized abnormalities (33). We also found that PCC connectivity with other regions, such as the left postcentral gyrus and right anterior insula, may actually be associated with rumination, rather than maladaptive SFA. These associations need to be confirmed in larger studies. Together, our findings suggest that PCC connectivity with the right superior parietal lobule, in particular, may reflect a specific neural signature of maladaptive SFA.

Our findings also contribute to a growing literature on abnormal patterns of connectivity in anxiety and obsessive-compulsive related disorders between regions within the DMN and other large-scale brain networks, such as the salience network (SN) and frontoparietal network (FPN) (25). For example, one study showed that individuals with obsessive-compulsive disorder displayed reduced anticorrelations between the DMN and the anterior insula (62), whereas another study found increased DMN connectivity with the insula (63). The anterior insula is a major node of the SN involved in processing interoceptive information, as well as detecting salient stimuli and switching between externally-oriented attention networks and internally-oriented self-referential networks to guide behavior (56-57). In our study, reduced PCC-anterior insula connectivity was not linked to maladaptive SFA. However, it may reflect abnormal interactions between the DMN and SN attributable to heightened anxiety sensitivity and interoceptive awareness to the physical sensations of anxiety, which has been described in SAD (36), but less well-studied in BDD. Part of the reason for inconsistent findings in studies examining DMN connectivity in anxiety and obsessive-compulsive related disorders (and perhaps why we did not confirm our original group-related hypothesis) is that the traditional case-control approach is limited by its ability to address comorbidities and other patient-related effects of an illness that are not present in controls. It is therefore important for future research conducting case-control comparisons of DMN connectivity to examine relationships between connectivity and clinical variables to better discern neural phenotypes associated with disorders.

A strength of our approach was the transdiagnostic examination of maladaptive SFA, which has implications for a common mechanism underlying two seemingly disparate psychiatric disorders, as well as the possibility of reclassification of these disorders in the DSM. SAD and BDD are currently classified as an anxiety disorder and an obsessive-compulsive spectrum disorder, respectively, in DSM-5. By enrolling individuals with SAD and BDD with similar levels of maladaptive SFA (as well as depression severity and insight), we identified a common pattern of dysconnectivity (between PCC and right superior parietal lobule) that correlated with Public SCS-R scores. This finding opens the possibility for engaging a common target in existing and new interventions for SAD and BDD.

Our study had some limitations. First, our Public SCS-R measure of trait maladaptive SFA was a subscale measure of trait self-consciousness that has not been well-studied in clinical populations. Despite the fact that there were only 7 items in this scale and we employed narrow selection criteria based on scores from this scale, there was enough variability in both the patient and control groups on this measure to demonstrate significant associations with our neural measures. More research is needed to examine the construct validity of maladaptive SFA as distinct from related constructs such as rumination and repetitive negative thinking, as well as its reliability as a stable trait-like construct. Second, the correlation between maladaptive SFA and PCC-right superior parietal lobule connectivity was unexpected and therefore requires replication. Third, we only examined connectivity with the PCC as a commonly studied seed region of the DMN; however, future studies could examine other nodes of the DMN using a similar seed-based approach, or in a multivariate manner, such as with independent components analysis. Lastly, our sample composition limits the generalizability of our findings to the general population, as our sample comprised of predominantly Caucasian, highly-educated, young, female participants. Studies have highlighted the problematic consequences of non-representative sampling on findings of brain structure and function (64-65), which underscore the need to broaden recruitment efforts and develop stronger community partnerships to capture the full dimension of variability in neural measures in clinical neuroscience studies.

Limitations notwithstanding, our study is the first to demonstrate the neural correlates of maladaptive SFA in a transdiagnostic sample. Specifically, PCC connectivity with the right superior parietal lobule may index maladaptive SFA across SAD and BDD, two disorders that are classified in separate categories in the DSM-5. Our findings lend support for a transdiagnostic conceptualization of maladaptive SFA as a cognitive dimension along the broad anxiety spectrum, represented by disruptions between large-scale brain networks involved in mediating internal and external attention. Our study provides the foundational basis for further research on the neural correlates of maladaptive SFA as a predictor of treatment response and potential novel target for intervention.

## Data Availability

The data that support the findings of this study are available from the corresponding author (A.F.) upon reasonable request.

## Acknowledgments and Disclosures

This work was supported by a NIMH Career Development Award K23 MH109593 to A.F. B.B. is supported by K01 MH114012. J.F. is supported by R01MH121520 (Feusner) and R21MH110865 (Feusner). S.W. has received research grant awards from Koa Health, royalty payments from New Harbinger Publications, Guilford Publications, Oxford University Press, and Springer, honorarium payments from the International OCD Foundation, Elsevier, Tourette Association of America and the Centers for Disease Control and Prevention, and Brattleboro Retreat, travel reimbursement from the International OCD Foundation and Brattleboro Retreat, as well as payment for serving on the Scientific Advisory Board of One-Mind PsyberGuide. No other co-authors have any competing interests to disclose.

The authors would like to acknowledge Dr. Mark Vangel for statistical assistance, Dr. Susanne Hoeppner for database management, and Rachel Porth, Jin Shin, and Lucas Petre for assistance with data acquisition.

