## Supplementary Information for "Maladaptive Self-Focused Attention and Default Mode Network Connectivity: A Transdiagnostic Investigation Across Social Anxiety and Body Dysmorphic Disorders"

Table S1. Psychiatric Comorbidities of Clinical Sample

Table S2. Associations Between DMN Functional Connectivity and Clinical Measures in Full Sample

Table S3. Clusters Showing Significant Group Differences in Functional Connectivity with the Posterior Cingulate Cortex (PCC) While Controlling for Medication Status

Table S1. Psychiatric comorbidities of clinical sample

| **Current Psychiatric Comorbidities** | **BDD (*n* = 15)**  **n (%)** | **SAD (*n* = 15)**  **n (%)** |
| --- | --- | --- |
| Social Anxiety Disorder | 8 (53.3) | --- |
| Body Dysmorphic Disorder | --- | 2 (13.3) |
| Generalized Anxiety Disorder | 3 (20.0) | 4 (26.7) |
| Major Depressive Disorder | 4 (26.7) | 3 (20.0) |
| Persistent Depressive Disorder | 2 (13.3) | 1 (6.7) |
| Panic Disorder | 2(13.3) | 1 (6.7) |
| Binge Eating Disorder | 0 (0.0) | 1 (6.7) |
| Feeding and Eating Disorder | 1 (6.7) | 2 (13.3) |
| Obsessive Compulsive Disorder | 2 (13.3) | 1 (6.7) |
| Agoraphobia | 0 (0.0) | 2 (13.3) |
| Specific Phobia | 2 (13.3) | 1 (6.7) |
| Attention Deficit Hyperactivity Disorder | 3 (20.0) | 0 (0.0) |
| Premenstrual Depressive Disorder | 0 (0.0) | 1 (6.7) |
| Posttraumatic Stress Disorder | 1 (6.7) | 1 (6.7) |

Table S2. Associations Between DMN Functional Connectivity and Clinical Measures

|  | 1 | 2 | 3 | 4 | 5 | 6 | 7 | 8 | 9 | 10 | 11 |
| --- | --- | --- | --- | --- | --- | --- | --- | --- | --- | --- | --- |
| 1. PCC-right CC | 1 | .562*** | .557*** | .596*** | .381* | .024 | -.198 | -.398 | -.378 | -.178 | -.232 |
| 2. PCC-right SPL | .555** | 1 | .257 | .868*** | .253 | .074 | -.573*** | -.417 | -.049 | -.150 | -.133 |
| 3. PCC-left PCG | .548** | .627*** | 1 | .324 | .507** | .397* | .034 | -.552* | -.439 | -.405* | -.433* |
| 4. PCC-left SPL | .638*** | .623*** | .664*** | 1 | .229 | .085 | -.530** | -.464 | -.345 | -.265 | -.198 |
| 5. PCC-right INS | .321 | .264 | .147 | .173 | 1 | .470** | -.017 | -.116 | -.615* | -.280 | -.402* |
| 6. PCC-left INS | .499** | .660*** | .503** | .521** | .470* | 1 | -.018 | -.127 | -.182 | -.242 | -.259 |
| 7. Public SCS-R | -.002 | -.048 | .131 | -.038 | -.104 | .126 | 1 | .378 | .354 | .135 | .095 |
| 8. LSAS | ^a^ | ^a^ | ^a^ | ^a^ | ^a^ | ^a^ | ^a^ | 1 | ^b^ | .482 | .333 |
| 9. BDD-YBOCS | ^a^ | ^a^ | ^a^ | ^a^ | ^a^ | ^a^ | ^a^ | ^a^ | 1 | .689** | .585* |
| 10. BDI-II | .219 | .284 | .265 | .256 | .053 | .198 | .038 | ^a^ | ^a^ | 1 | .681*** |
| 11. RRS | .042 | .059 | -.067 | .014 | .258 | .050 | -.123 | ^a^ | ^a^ | .271 | 1 |

*Notes*. Upper diagonal reflects patients; lower diagonal (shaded) reflects controls. PCC = posterior cingulate cortex; CC = cingulate cortex; SPL = superior parietal lobule; PCG = postcentral gyrus; INS = insula; SCS-R = Self-Consciousness Scale- Revised; LSAS = Liebowitz Social Anxiety Scale; BDD-YBOCS = Yale-Brown Obsessive-Compulsive Scale Modified for BDD. BDI-II = Beck Depression Inventory-II. RRS = Ruminative Responses Scale. ***p≤.001, **p≤.01, *p≤.05. ^a^LSAS and BDD-YBOCS were not administered to healthy controls. ^b^LSAS and BDD-YBOCS correlations not available since LSAS was only administered to patients with primary SAD and BDD-YBOCS was only administered to patients with primary BDD.

Table S3. Clusters Showing Significant Group Differences in Functional Connectivity with the Posterior Cingulate Cortex (PCC) While Controlling for Medication Status

| Region | Voxel size | MNI Coordinates | | | BA | t(56) |
| --- | --- | --- | --- | --- | --- | --- |
|  |  | x | y | z |  |  |
| R insula/central operculum | 414 | 38 | -2 | 4 | - | 4.27 |
| R postcentral gyrus/superior parietal lobule | 354 | 20 | -42 | 80 | - | 4.10 |
| R cingulate cortex | 338 | 8 | 2 | 44 | 24 | 4.22 |
| L postcentral gyrus | 319 | -42 | -18 | 28 | 2 | 4.65 |
